# Prevalence of different sleep problems in patients with respiratory diseases presented to a respiratory outdoor clinic: A Descriptive analysis

**DOI:** 10.1101/2021.06.17.21258633

**Authors:** Arup Kumar Haldar, Arpita Halder Chatterjee

**Author notes:** (Corresponding Author) Mail.

## Abstract

**Introduction:** A patient with compromised respiratory system due to some diseases, may have disturbed sleep to a great extent. It has been proved in various community based epidemiologic studies. Though most of such studies available had concentrated on single respiratory disease like COPD or Asthma in the community. But such studies are hardly available for patients attending a respiratory clinic, where all respiratory ailments were evaluated with concurrent sleep problems. The present study is one such.

**Methods:** Total 163 patients were screened and among them 100 were selected as the study group. The patients were enquired with a standard questionnaire provided to them. In addition, they were given seperate questionnaire as STOP BANG, Insomnia Severity Index Score, RLS questionnaire. Those persons with STOP BANG > 5, were also advised for a Polysomnography (PSG), if not already done. Descriptive statistical analysis has been carried out in the present study.

**Results:** The present study is a cross-sectional depiction of relationships between various respiratory diseases and sleep problems. As per this study, most patients were asthmatic and commonest sleep problem was OSA. A subgroup analysis was done to determine the significance of difference of various parameters between the three groups of asthmatic patients, patients having COPD and the ‘Other’ group.

**Discussion:** Subjective sleep problems were significantly more in Asthma group than COPD group (p<0.0362). The mean STOP BANG was more in COPD group than the Asthma group (p<0.0301). Though OSA was the commonest sleep problem between the three groups, but the prevalence was not statistically significant between them. More patients in the COPD group had insomnia, but it was not statistically significantly more than in Asthma group. Sleeping pill use was significantly more in COPD group than the Asthma group (p<0.0039).

**Conclusion:** Sleep problems are common in patients with respiratory diseases and OSA is the most common problem according to the present study. Asthma patients had more subjective sleep problems. Sleeping pill use is more common in COPD patients instead of having less subjective sleep complaints than asthma patients.

## Introduction

The quality and quantity of sleep are the determinants of quality of life. But performance of respiratory system is not at its best during sleep. So a patient with compromised respiratory system due to some diseases, may have disturbed sleep to a great extent. In the Tuscan epidemiologic study it was found that 53% of patients with COPD experience difficulties in initiating or maintaining sleep and 26% complaints of excessive daytime sleepiness (EDS), compared to 36% and 11%, respectively, in age matched controls having no respiratory diseases [1]. Similarly Asthma patients are also prone to various sleep problems. In an analysis of 225 patients from the SARP Cohort, 40% of severe Asthmatic complaints of EDS, and 31% had a elevated Epworth Sleepiness Score (EPSS) [2]. Inspite of such wide prevalence of sleep problems in Respiratory patients we often do not take the sleep history from the patient. In 2013, Price et al showed that, although 78% patients reported nighttime symptoms, only 67% of clinicians reported that their patients were bothered at night [3]. In view of all these, the present study was done in a Respiratory outdoor clinic to evaluate different sleep problems in a small group of patients presenting with various respiratory ailments.

## Methods

The study was carried out in 2019 in a Respiratory Clinic in Kolkata (February to June). All patients coming for some respiratory consultations were requested to provide the data required for the study. The inclusion criteria were-

1. Adult patients > 18 Years
2. Not terminally ill
3. Follow up data available for atleast 6 months

The patients were enquired with a standard questionnaire provided to them. In addition, they were given seperate questionnaire as STOP BANG [4], Insomnia Severity Index Score [5], RLS questionnaire [6]. Those persons with STOP BANG > 5, were also advised for a Polysomnography (PSG) [7], if not already done. Only those patients having a PSG documented for OSA presence were tabulated as confirmed OSA.

Total 163 patients were screened and among them 100 were selected as the study group, for whom we are able to gather all the required informations. Descriptive statistical analysis has been carried out in the present study. Results on continuous measurements are presented on Mean ±SD and results on categorical measurements are presented in Number (%). Significance is assessed at a level of 5%. Statistical software: MedCalc Software [8].

## Results

The total number of patients in this study was 100. The distribution of patients according to the diagnosis were asthma =45, COPD =24, Tuberculosis =4, acute bronchitis=2, Respiratory tract infection= 20, lung cancer=1, interstitial lung disease =2, bronchiectasis=1, GERD=1. The most common diagnosis in this population was asthma, then followed by COPD, followed by acute respiratory tract infections. The characteristics of each of these group is given in Table 1. Those patients not having either Asthma or COPD were clubbed together as ‘Other’ group.

**Table 1.**
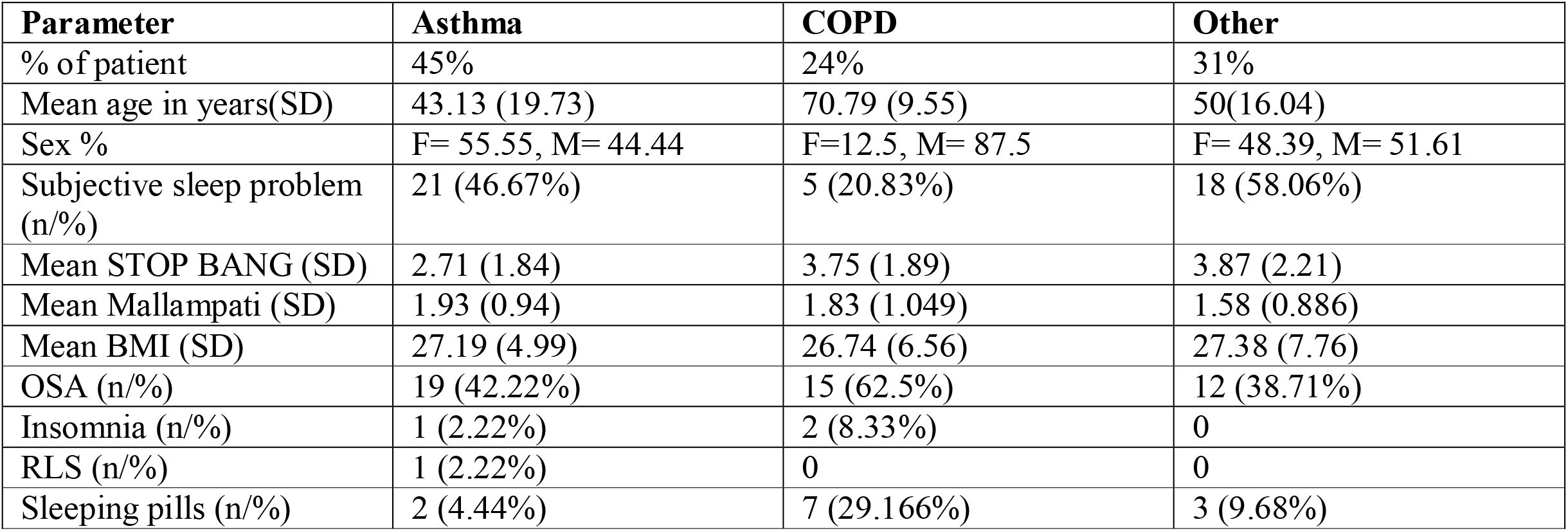

The above table is self explanatory. Now a subgroup analysis were done between these three groups to determine the level of significance of the observations between them. The Table 2 is related to that calculation.

**Table2.**

| Parameter | Asthma | COPD | Other | Difference | Standard error | 95% CI | t statistics | Chi-squared | DF | P value |
| --- | --- | --- | --- | --- | --- | --- | --- | --- | --- | --- |
| **Mean age in years** | **43.13** | **70.79** |  | **27.66** | **4.28** | **19.11to 36.20** | **6.46** |  | **67** | **<0.001** |
| Mean age in years | 43.13 |  | 50.16 | 7.03 | 4.27 | -1.49 to 15.55 | 1.644 |  | 74 | 0.1045 |
| **Subjective sleep problems** | **46.67%** | **20.83%** |  | **25.84%** |  | **1.88 to 44.21%** |  | **4.386** | **1** | **0.0362** |
| Subjective sleep problems | 46.67% |  | 58.06 % | 11.39% |  | -14.96 to 35.09% |  | 0.657 | 1 | 0.4178 |
| **Mean STOP-BANG** | **2.71** | **3.75** |  | **1.040** | **0.469** | **0.103 to 1.977** | **2.215** |  | **67** | **0.0301** |
| Mean STOP-BANG |  | 3.75 | 3.87 | -0.120 | 0.565 | -1.25 to 1.01% | -0.212 |  | 53 | 0.8326 |
| **Mean STOP-BANG** | **2.71** |  | **3.87** | **-1.160** | **0.466** | **-2.089 to -0.231** | **-2.487** |  | **74** | **0.0151** |
| Mean Mallampati | 1.93 | 1.83 |  | -0.100 | 0.247 | -0.593 to 0.394 | -0.404 |  | 67 | 0.6874 |
| Mean Mallampati | 1.93 |  | 1.58 | -0.350 | 0.214 | -0.777 to 0.077 | -1.633 |  | 74 | 0.1068 |
| Mean Mallampati |  | 1.83 | 1.58 | -0.250 | 0.261 | -0.773 to 0.274 | -0.958 |  | 53 | 0.3426 |
| Mean BMI | 27.19 | 26.74 |  | -0.450 | 1.41 | -3.26 to 2.36 | -0.319 |  | 67 | 0.7506 |
| Mean BMI | 27.19 |  | 27.38 | 0.190 | 1.46 | -2.72 to 3.10 | 0.130 |  | 74 | 0.8969 |
| Mean BMI |  | 26.74 | 27.38 | 0.64 | 1.97 | -3.32 to 4.60 | 0.324 |  | 53 | 0.7472 |
| OSA | 42.22% | 62.5% |  | 20.28% |  | -4.24 to 41.32% |  | 2.538 | 1 | 0.1111 |
| OSA | 42.22% |  | 38.70 % | 3.52% |  | -18.40 to 24.35% |  | 0.093 | 1 | 0.760 |
| OSA |  | 62.5% | 38.70 % | 23.8% |  | -2.59% to 45.96% |  | 3.010 | 1 | 0.0827 |
| Insomnia | 2.22% | 8.33% |  | 6.11% |  | -5 to 23.72% |  | 1.385 | 1 | 0.2392 |
| **Sleeping pills** | **4.44%** | **29.16%** |  | **24.72%** |  | **7.089 to 44.977%** |  | **8.313** | **1** | **0.0039** |
| Sleeping pills | 4.44% |  | 9.68% | 5.24% |  | -6.92% to 20.799% |  | 0.809 | 1 | 0.3683 |
| Sleeping pills |  | 29.16% | 9.68% | 19.48% |  | -1.37 to 40.459% |  | 3.388 | 1 | 0.0657 |

The observations which achieved the level of significance are written in bold.

## Discussion

The present study is a cross-sectional depiction of relationships between various respiratory diseases and sleep problems. As per Table 1, most patients were asthmatic and commonest sleep problem was OSA. A subgroup analysis was done in Table 2 to determine the significance of difference of various parameters between the three groups of asthmatic patients, patients having COPD and the ‘Other’ group.

As per the result the mean ages of Asthma and COPD patients were significantly different, as expected. Subjective sleep problems were significantly more in Asthma group than COPD group. The mean STOP BANG was more in COPD group than the Asthma group. It may be explained by age related propensity to develop OSA [9]. Mean STOP BANG was also significantly more in ‘Other’ group than Asthma group, which is difficult to explain as heterogeneity of age in ‘Other’ group. The difference between MALLAMPATI and BMI were not significant in between the groups. Though OSA was the commonest sleep problem between the three groups, but the prevalence was not statistically significant between them. More patients in the COPD group had insomnia, but it was not statistically significantly more than in Asthma group. Sleeping pill use significantly more in COPD group than the Asthma group.

If results are pulled together, this study shows-

1. The commonest sleep problem in Respiratory practice is OSA
2. Subjective sleep problems are more in Asthma patients.
3. The COPD group has a increased STOP BANG, though the prevalence of OSA was not more than Asthma group or from ‘Other’ respiratory patients. This may be related to a Subjective error in calculation of STOP BANG, or may be due to increased comorbidities, like Hypertension with increasing age.
4. Though the incidence of insomnia was not statistically more in COPD, still the use of sleeping pill was significantly more in COPD, inspite of having lesser number of subjective sleep complaints than Asthma patients. It is difficult to ascertain whether this is due to the effects of sleeping pills or not.

Sleep problems in respiratory patients are very common and large community based epidemiologic studies are available. But such studies are lacking based on a respiratory clinic.

Most of such studies available had concentrated on single respiratory disease like COPD or Asthma. Few of such studies in COPD had reported a wide prevalence of different sleep problems as follows- : RLS 29-36% patients [10-12], overlap syndrome or comorbid OSA, 11–16% patients [13], Insomnia 27-53% patients [14], in patients with moderate-to-severe COPD. Though based on these studies insomnia or RLS were quite prevalent in COPD patients, but the present study was unable to detect such relationship. It may be due to subjective measure of RLS or Insomnia, which are always subjected to bias. Still the present study revealed a good number of COPD patients use sleeping pills, probably to control their sleep problems. Such therapy may be dangerous in subgroup of COPD patients with nocturnal or daytime hypoventilation [15]. The weakness of this study was a low sample size. Beside that the results could not be extrapolated to general population as a special group of people were screened, who presented to respiratory clinic. We need a prospective cohort study for definite evaluation of association between various respiratory diseases and sleep problems in them.

## Conclusion

Sleep problems are common in patients with respiratory diseases and OSA is the most common problem according to the present study. Asthma patients had more subjective sleep problems. Sleeping pill use is more common in COPD patients instead of having less subjective sleep complaints than asthma patients. All respiratory physician must include a sleep history for evaluation of such problems and their remedy.

## Data Availability

All data are available

---

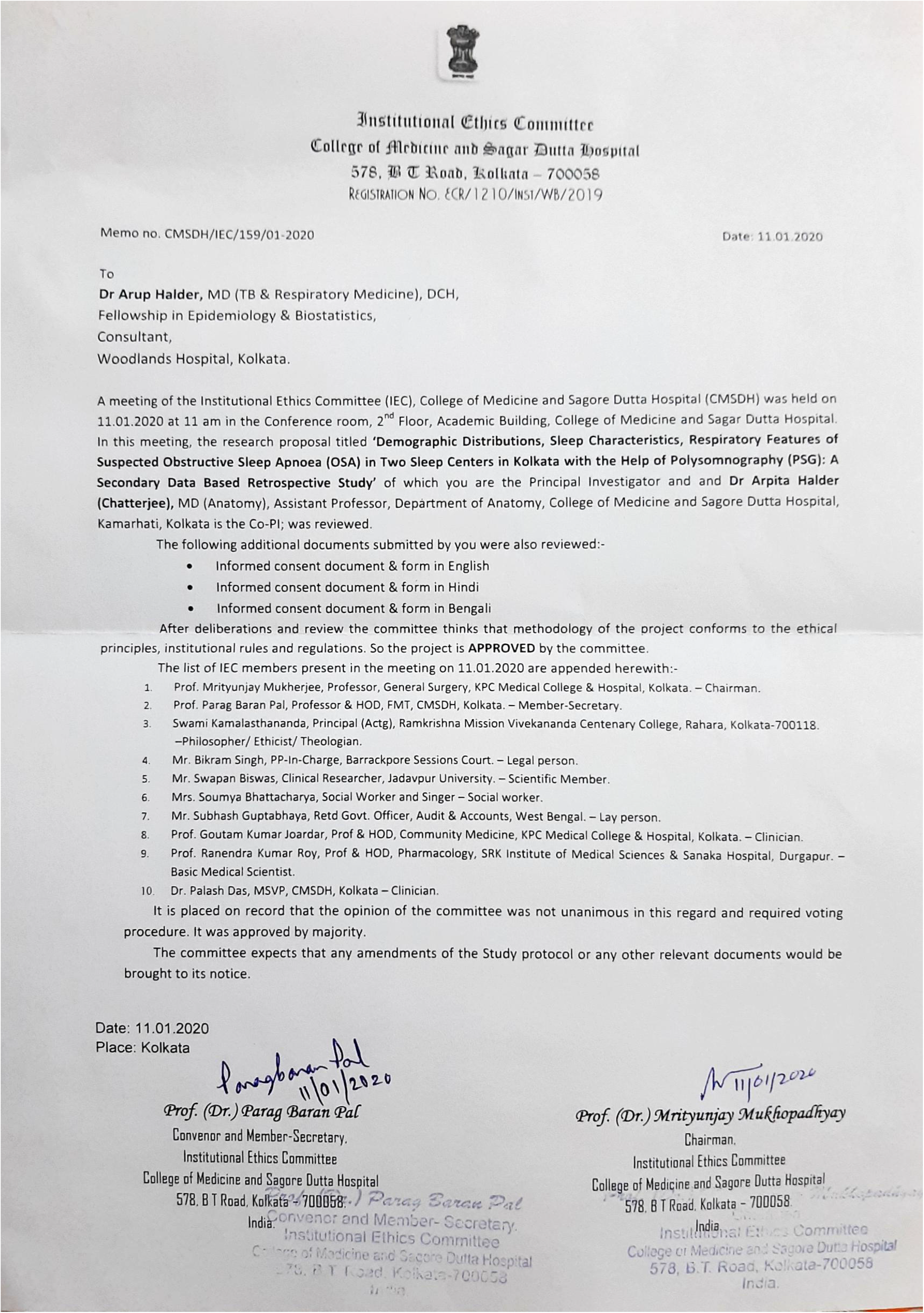

## Notes

### Competing Interest Statement

The authors have declared no competing interest.

### Funding Statement

Nil

### Author Declarations

Ethical Committee Clearance taken and submitted in the required section as an upload. Registration number of the Ethical committeeECR/1210/WST/WB/2019, Institution Ethics Committee, Collefe of Medicine and Sagar Duuta Hospital, 578 BT Road, Kolkata 700058, West Bengal, India.

## References

1. Mary E. Klink, Russell Dodge, Stuart F. Quan, The Relation of Sleep Complaints to Respiratory Symptoms in a General Population, Chest, Volume 105, Issue 1, 1994, Pages 151154,ISSN 0012-3692, https://doi.org/10.1378/chest.105.1.151. (https://www.sciencedirect.com/science/article/pii/S0012369215438358)

2. Mihaela Teodorescu, Oleg Broytman, Douglas Curran-Everett, Ronald L. Sorkness, Gina Crisafi, Eugene R. Bleecker, Serpil Erzurum, Benjamin M. Gaston, Sally E. Wenzel, Nizar N. Jarjour, Obstructive Sleep Apnea Risk, Asthma Burden, and Lower Airway Inflammation in Adults in the Severe Asthma Research Program (SARP) II, The Journal of Allergy and Clinical Immunology: In Practice, Volume 3, Issue 4, 2015, Pages 566-575.e1, ISSN 2213-2198, https://doi.org/10.1016/j.jaip.2015.04.002. (https://www.sciencedirect.com/science/article/pii/S2213219815001762)

3. Price D, Small M, Milligan G, Higgins V, Gil EG, Estruch J. The prevalence and impact of night-time symptoms in COPD-a real-world study in five European countries. Int J Chron Obstruct Pulmon Dis. 2013;8:595–603.

4. Frances Chung, Hairil R. Abdullah, Pu Liao, STOP-Bang Questionnaire: A Practical Approach to Screen for Obstructive Sleep Apnea, Chest, Volume 149, Issue 3, 2016, Pages 631–638, ISSN 0012-3692, https://doi.org/10.1378/chest.15-0903. (https://www.sciencedirect.com/science/article/pii/S0012369215000185)

5. Célyne H Bastien, Annie Vallières, Charles M Morin, Validation of the Insomnia Severity Index as an outcome measure for insomnia research,Sleep Medicine, Volume 2, Issue 4, 2001, Pages 297–307,ISSN 1389-9457, https://doi.org/10.1016/S1389-9457(00)00065-4. (https://www.sciencedirect.com/science/article/pii/S1389945700000654)

6. Richard P. Allen, Brendan J. Burchell, Ben MacDonald, Wayne A. Hening, Christopher J. Earley, Validation of the self-completed Cambridge-Hopkins questionnaire (CH-RLSq) for ascertainment of restless legs syndrome (RLS) in a population survey,Sleep Medicine, Volume 10, Issue 10, 2009, Pages 1097–1100, ISSN 1389-9457, https://doi.org/10.1016/j.sleep.2008.10.007. (https://www.sciencedirect.com/science/article/pii/S1389945708003535)

7. N.J Douglas, S Thomas, M.A Jan, Clinical value of polysomnography, The Lancet, Volume 339, Issue 8789, 1992, Pages 347–350, ISSN 0140-6736, https://doi.org/10.1016/0140-6736(92)91660-Z. (https://www.sciencedirect.com/science/article/pii/014067369291660Z)

8. MedCalc Software Ltd. Comparison of means calculator. https://www.medcalc.org/calc/comparison_of_means.php (Version 20.006; accessed June 9, 2021)

9. Janssens, JP., Pautex, S., Hilleret, H. et al. Sleep disordered breathing in the elderly. Aging Clin Exp Res 12, 417–429 (2000). https://doi.org/10.1007/BF03339872)

10. Dipti Gothi. Sleep disorders in chronic obstructive pulmonary disease, Indian J Sleep Med 2015; 10.1, 11-21, DOI No:10.5958/0974-0155.2015.00002.9

11. Lo Coco D, Mattaliano A, Lo Coco A, Randisi B. Increased frequency of restless legs syndrome in chronic obstructive pulmonary disease patients. Sleep Med. 2009;10:572–576.

12. Cavalcante AG, de Bruin PF, de Bruin VM, et al. Restless legs syndrome, sleep impairment, and fatigue in chronic obstructive pulmonary disease. Sleep Med. 2012;13: 842–847

13. Budhiraja R, Parthasarathy S, Budhiraja P, Habib MP, Wendel C, Quan SF. Insomnia in patients with COPD. Sleep. 2012;35:369–75

14. Lacasse Y, Sériès F, Vujovic-Zotovic N, et al. Evaluating nocturnal oxygen desaturation in COPD revised. Respir Med. 2011;105:1331–1337.

15. George, C.F., Bayliff, C.D. Management of Insomnia in Patients With Chronic Obstructive Pulmonary Disease. Drugs 63, 379–387 (2003). https://doi.org/10.2165/00003495-200363040-00004)

